# Hydrocortisone for the prevention of postoperative delirium in digestive surgery (HyPOD): Study protocol for a multicentre randomized, blind, placebo-controlled trial

**DOI:** 10.1101/2025.05.06.25327048

**Authors:** Tsukasa Aritake, Yukihiro Yokoyama, Masahiko Ando, Yachiyo Kuwatsuka, Yumi Suzuki, Yuji Shingu, Koichi Akiyama, Tomoki Ebata

## Abstract

**Introduction:** Postoperative delirium (POD) is a significant complication in digestive surgery, potentially increasing hospital stays, costs, and mortality. Corticoids, such as dexamethasone and methylprednisolone, have been reported to reduce POD. However, the efficacy of hydrocortisone (HC) remains unexplored. This trial aims to evaluate HC’s effectiveness in preventing POD in surgical patients.

**Methods and analysis:** A multicentre (three hospitals), randomized, blinded (patient-blinded, surgeon-blinded, anaesthesiologist-blinded, and monitor-blinded), placebo-controlled trial of HC for the prevention of POD in patients undergoing digestive surgery was started in June 2024. Patients undergoing digestive surgery are randomly assigned to the HC or placebo group. The stratification factors are age, sex, disease stage, and procedure type. Before skin incision, the participants in the HC group are administered 500 mg HC intravenously. The participants in the placebo group are administered the same volume of saline that is indistinguishable from the HC. The primary outcome is the presence of POD from postoperative Days 1 to 3, which is assessed using the Confusion Assessment Method-ICU (CAM-ICU). The secondary outcomes are changes in serum sodium levels, postoperative mortality, length of hospital stay, functional independence measures, and complications within 28 days. To date, 61 patients of a planned 180 have been enrolled in the study.

**Ethics and dissemination:** This protocol was approved by the Nagoya University Clinical Research Review Board and was registered with the Japan Registry of Clinical Trials on 24 May 2024. The results of this trial will be disseminated through peer-reviewed journals.

**Trial registration number:** jRCTs041240032

**Strengths and limitations of this study:**

- This randomized controlled trial (RCT) is the first study to answer the clinical question of whether the preoperative use of hydrocortisone (HC) in digestive surgery is beneficial.
- A multicentre (three hospitals), double-blinded (patient-, surgeon-, anaesthesiologist-, and monitor-blinded) placebo-controlled RCT is the optimal study design for addressing the clinical question above.
- A loading dose of HC (500 mg), which is simple and easy for anaesthesiologists to administer, is administered intravenously over 15–30 min just before surgery.
- The study inclusion criteria are extensive, and the exclusion criteria are limited; therefore, this protocol will have broad applicability if the trial results are positive.
- On the basis of previous data regarding postoperative delirium (POD) from the National Center for Geriatrics and Gerontology (NCGG), the necessary sample size is estimated to be 180 patients; however, the incidence of POD tends to vary widely among institutions, and the risk of underpowered statistics is not negligible.

## Introduction

Postoperative delirium (POD) is among the most important complications in the field of gastrointestinal surgery. Patients who develop POD tend to have longer hospital stays, higher medical costs, cognitive dysfunction, and increased mortality (1–3).

Recently, the effectiveness of pharmacological treatments, particularly those targeting insomnia, and nonpharmacological therapies, such as perioperative rehabilitation, have become well established in the management of POD (4–7). However, the incidence of POD remains high after gastrointestinal surgery, with a reported incidence of approximately 24% (8), whereas it has reached as high as 39.4% at the National Center for Geriatrics and Gerontology (NCGG).

Corticoids, steroid hormones from the adrenal cortex, reduce inflammatory responses in postoperative patients (9–11). Four recent randomized controlled trials (RCTs) reported that steroids with minimal mineralocorticoid effects may prevent POD (12–15); two studies treated laparoscopic gastrointestinal surgery and open hepatectomy in each (16, 17) (Table 1). Considering that hyponatremia is a risk factor for POD (18–20), mineralocorticoids, which increase sodium and water reabsorption in the distal tubules and collecting ducts of the kidney, may help prevent POD as well as postoperative hyponatremia. However, there have been no interventional studies on the efficacy of steroid preparations with a representative mineralocorticoid, hydrocortisone (HC), in reducing POD.

**Table 1.** List of previous RCTs on the ability of corticoids to prevent postoperative delirium. *Kluger et al. reported a statistically significant difference (P value=0.010) in the severity, but not the incidence, of delirium.

| year | Authors | department | number of patients | drug | postoperative delirium placebo vs. intervention | P value |
| --- | --- | --- | --- | --- | --- | --- |
| 2018 | Clemmesen et al<sup>12</sup> | orthopedics | 117 | methylprednisolone | 32.8% vs 16.9% | 0.048 |
| 2020 | Zhang et al<sup>13</sup> | orthopedics | 218 | dexamethasone | 30.6% vs 18.2% | 0.033 |
| 2021 | Xiang et al<sup>16</sup> | digestive surgery | 168 | methylprednisolone | 23.8% vs 10.7% | 0.03 |
| 2021 | Kluger et al<sup>14</sup> | orthopedics | 79 | dexamethasone | 23.1% vs 15.0% | 0.36* |
| 2022 | Awada et al<sup>17</sup> | digestive surgery | 53 | dexamethasone | 18.5% vs 0% | - |
| 2023 | Huang et al<sup>15</sup> | orthopedics | 160 | dexamethasone | 26.3% vs 11.3% | 0.015 |

### Study objectives

The key objective is to test our hypothesis that HC given preoperatively reduces POD.

## Methods and Analysis

### Study design and setting

HC for the prevention of POD in patients undergoing digestive surgery (HC-POD) is a multicentre, randomized, double-blind, placebo-controlled phase II trial. This study was conceived and initiated at Nagoya University Hospital, an academic referral hospital in Nagoya, Aichi, Japan. NCGG and Tsushima City Hospital are participating in this study. Patients are randomly assigned to receive either HC (intervention) or a placebo (control) just before surgery. In addition to the intervention, each institution’s protocols are followed in clinical practice. Permissive eligibility criteria, a simple study protocol, and limited data collection are being adopted to maintain a pragmatic study design and study feasibility. The study design is shown in Figure 1, and the timeline of assessment and follow-up is presented in Table 2. The final report of this study will follow the Consolidated Standards of Reporting Trials statement. This study protocol was written according to the Standard Protocol Items Recommendations for Interventional Trials guidelines (Supplemental Additional File 1).

**Figure 1.**
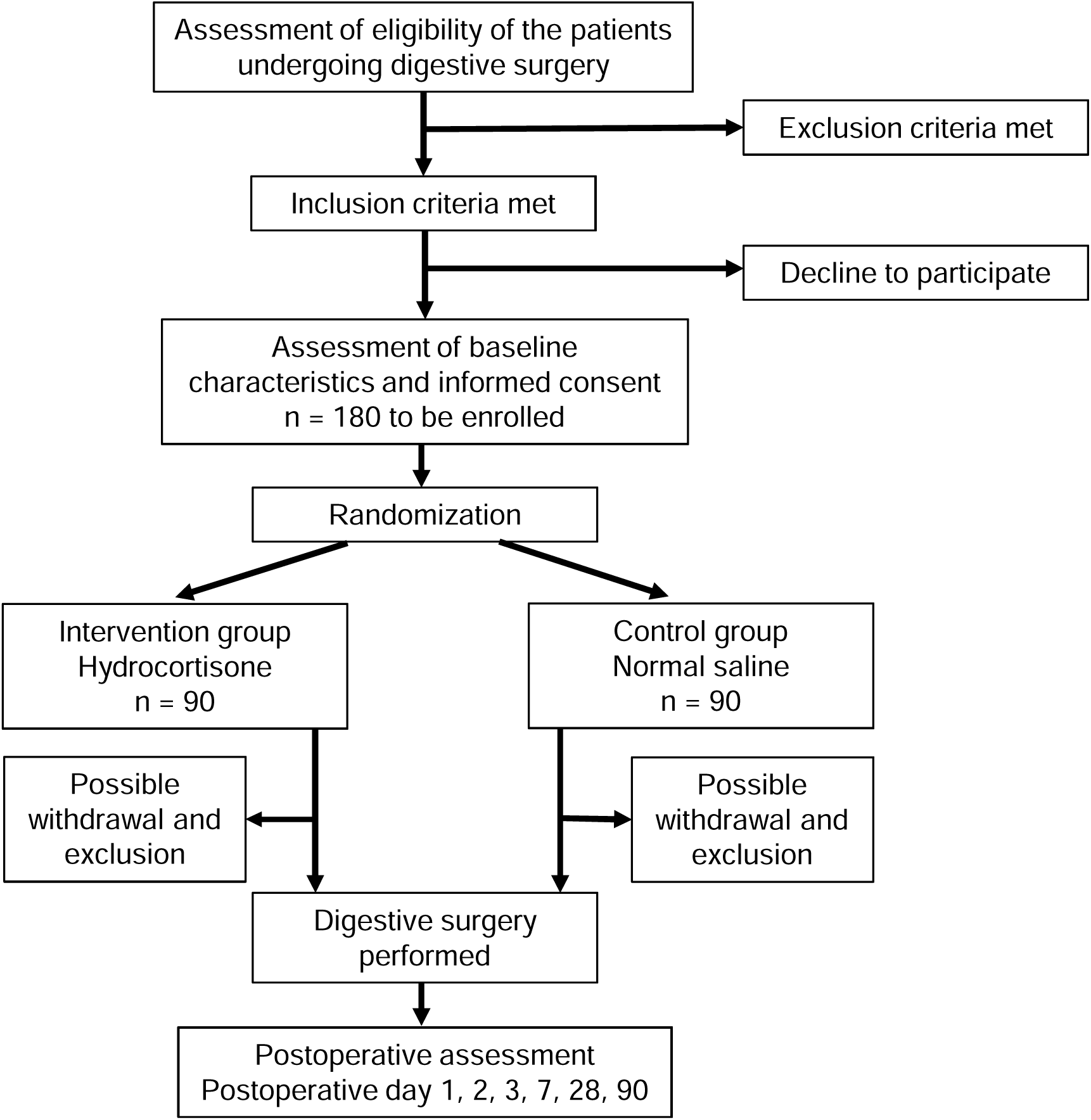
Study design.

**Table 2.**
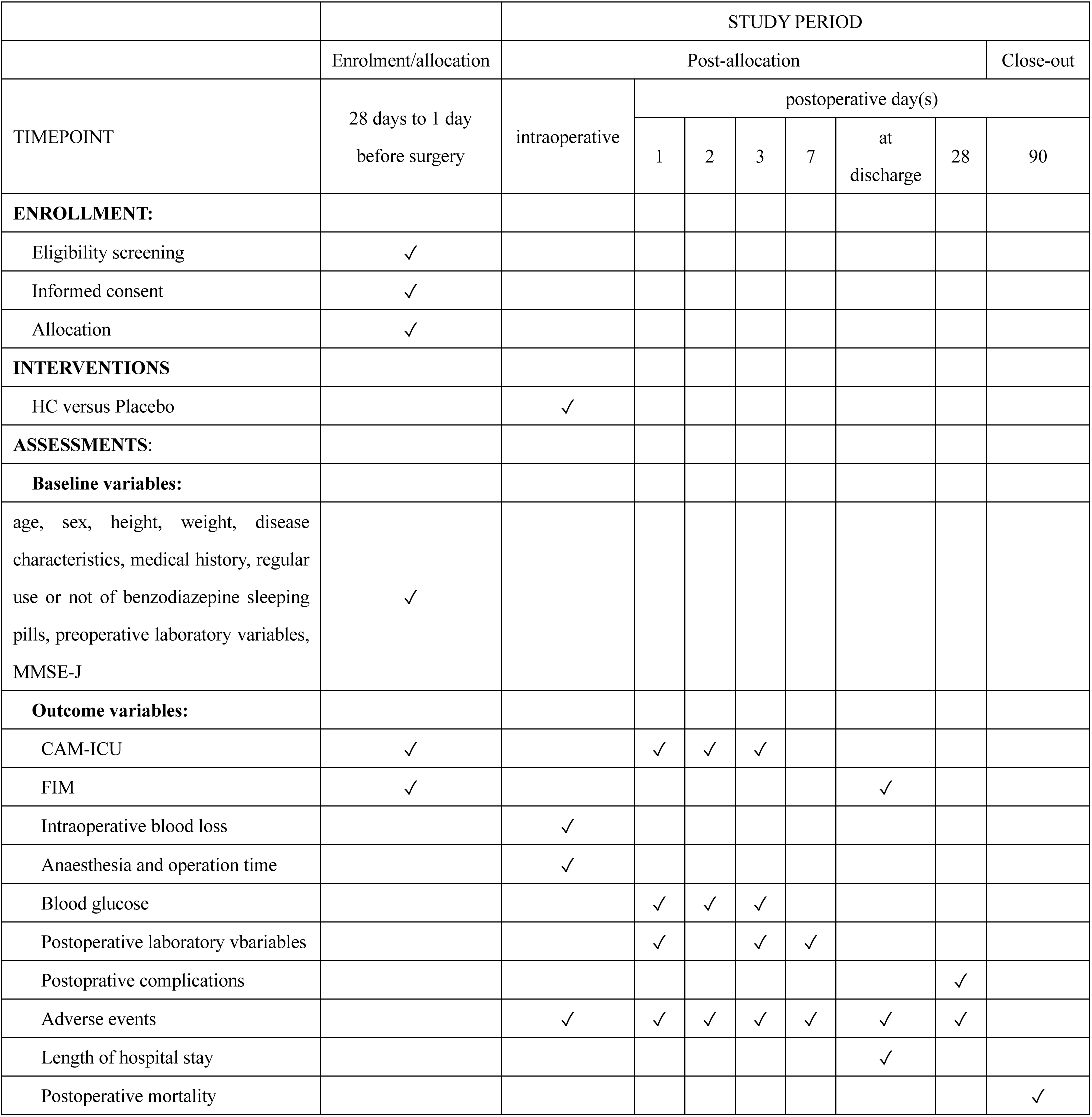
Assessment and follow-up timeline of the HC-POD study. HC, hydrocortisone; MMSE-J, Mini-Mental State Examination–Japanese; CAM-ICU, Confusion Assessment Method–Intensive Care Unit; FIM, Functional Independence Measure.

### Ethics, registration, and informed consent

This RCT will be conducted in accordance with the principles of the Declaration of Helsinki. In addition, because the prophylactic use of HC in this study is considered off-label in Japan, this study is considered a specified clinical trial as defined by the Clinical Trials Act published by the Ministry of Health, Labour and Welfare of Japan. The study protocol and consent forms were reviewed as a specified clinical trial and approved by the Nagoya University Clinical Research Review Board. This study was registered with the Japan Registry of Clinical Trials. Written informed consent is obtained from all participants.

### Patient inclusion criteria

Patients undergoing planned digestive surgery are eligible for the study. Eligible procedures include laparotomy, laparoscopic gastrointestinal surgery, and laparoscopic cholecystectomy for cholecystitis. Surgeries for inguinal hernia and laparoscopic cholecystectomy for noninflammatory gallbladder stone disease and gallbladder polyps are excluded because these procedures are expected to be less invasive with a short postoperative hospital stay after surgery. Eligible patients are those who are 65 years of age or older, of any sex, and who understand and agree with the concept of the study.

### Patient exclusion criteria

Patients are excluded from enrolment if any of the following criteria are met: (1) having an allergy to HC, (2) receiving systemic corticoids or immunosuppressive therapy, (3) having active peptic ulcer disease, (4) having insulin-dependent diabetes mellitus, (5) having a history of stroke within 3 months, (6) having no history of measles or varicella and having no prior measles or varicella vaccination, (7) having a history of hepatitis B or hepatitis C, (8) having severe communication disorders in vision or hearing, (9) having unstable mental status or psychiatric disorders, (10) currently participating in other intervention studies that may affect the results of this study, or (11) undergoing emergency surgery. These criteria are screened by investigators just before informed consent and enrolment.

### Trial interventions

Japanese-sourced HC (hydrocortisone made by Nichi-Iko Pharmaceutical Co., Ltd.) is used in this study. A loading dose of HC is administered intravenously at a dose of 500 mg (10 mL of HC diluted with 40 mL of 0.9% sodium chloride [normal saline]) over 15 to –30 min just before surgery. In the placebo group, the same amount of normal saline is administered intravenously at the same rate.

These drug (or placebo) administrations are monitored by surgeons, anaesthesiologists, and other medical staff members who are also blinded to the assignment.

### Rationale of the dose

Currently, no RCTs have examined the effect of HC on the prevention of POD. In the present study, we use a dose of 500 mg of HC because the potency of glucocorticoid action is nearly equivalent to the 2 mg/kg of methylprednisolone actually used in previous studies (16).

### Randomization and assignment

The enrolled patients are randomly assigned to the HC arm or placebo arm at a 1:1 ratio no earlier than 28 days before surgery. Investigators perform web-based online enrolment after obtaining written informed consent from the patients. Randomization is conducted using a stochastic minimization procedure stratified by study centre (three hospitals), sex, age (<75 years), disease (benign disease, low-stage malignant disease [≤cStage II], and high-stage malignant disease [≥cStage III]), and procedure (laparoscopic cholecystectomy, laparoscopic gastrointestinal surgery, or open abdominal surgery). Investigators are required to provide information on these five stratification factors with the data centre at the time of enrolment. An electronic data capture (EDC) system is used to perform the randomization and data collection. The allocation sequence and the EDC are generated and managed by the data centre at Nagoya University Hospital.

### Blinding and drug preparation

The result of the randomized assignment is sent to predetermined open-label pharmacists at each centre by e-mail from the data centre. The open-label pharmacists prepare the drug or placebo at the pharmacy department on the day of surgery. They cannot divulge allocation results unless a serious adverse event (SAE) occurs after the intervention and unblinding is requested. One ampoule of HC (500 mg/10 mL) is mixed with 40 mL of normal saline when the patients are assigned to the HC group, and only 50 mL of normal saline is prepared in the placebo group. Mixing by the open-label pharmacists is completely out of sight of the other staff members. Anaesthesiologists administer the prepared drug or placebo intravenously in the operating room. Study drugs are stored in a locked place at each centre’s pharmacy department and are managed only by the open-label pharmacists. This protocol achieves blinding of the patients, investigators, surgeons, anaesthesiologists, and other medical staff members except for the open-label pharmacists because the diluted HC is colourless and indistinguishable from the placebo. Members of the Data Safety and Monitoring Board (DSMB) are also blinded from the results of randomization, and only the statistical manager at the data centre has the right to access the results.

### Other treatments

Other procedures, drugs, instruments, and postoperative management are at the discretion of the surgeon and anaesthesiologist. Treatment of any complications that develop postoperatively is permissible. Corticoid medications other than the interventions used in this study are not allowed.

### Outcome measures

The primary outcome is the incidence of POD until postoperative Day 3. Delirium is measured twice a day (during the day and night shifts) from 1 to 3 days after surgery via the Confusion Assessment Method–Intensive Care Unit (CAM-ICU) (21). If the CAM-ICU is positive at least once, POD is present. The Richmond Agitation and Sedation Scale (RASS) is used to evaluate the sedation level of every patient before the CAM-ICU assessment (22), with patients scoring −4 or −5 excluded.

Secondary outcomes include changes in serum sodium levels on postoperative Days 1, 3, and 7; postoperative mortality at 90 days; length of hospital stay after surgery; and changes in FIM (functional independence measure) scores from admission to discharge. The FIM is a tool that quantifies the severity of disability in rehabilitation patients and consists of 18 items: 13 on motor functions and 5 on cognitive functions (23). Secondary outcomes also include blood glucose levels in the first 3 postoperative days and the incidence and severity of complications within 28 days after surgery. Complications include postoperative pancreatic fistula (POPF), haemorrhage associated with POPF, bile fistula, delayed gastric emptying, intra-abdominal abscess, wound infection, bowel obstruction, intestinal paralysis, anastomotic ulcer, sepsis, pneumonia, urinary tract infection, heart failure, acute kidney injury, hepatic failure, postoperative bleeding, thrombotic and embolic complications, seizures, and drug allergy, which are evaluated using the ClavienLDindo classification (24). SAEs are defined as any complication with a ClavienLDindo grade of 4a or higher.

### Data collection and follow-up

The case report forms in this study are constructed as web-based forms (from the EDC). The data are entered by blinded investigators before and after surgery (postoperative Days 1, 2, 3, 7, 28, and 90) and are fully supported by source documents. The baseline patient characteristics include age, sex, disease history, comorbidities, regular use of benzodiazepines for sleep induction, preoperative diagnosis, preoperative laboratory variables (white blood cell count, haemoglobin concentration, platelet count, serum albumin, serum creatinine, total serum bilirubin, and C-reactive protein), and preoperative urinary sodium levels. The CAM-ICU, Mini-Mental State Examination–Japanese (MMSE-J), and FIM at admission are also included. The data that will be captured and relevant to the surgery include the surgical procedure, anaesthesia time, surgery time, and intraoperative blood loss. Postoperative management is performed as per-standard care at each institution; however, blood glucose levels and CAM-ICU scores on the first three postoperative days and blood tests and urinary sodium levels on postoperative Days 1, 3, and 7 are required. Postoperative complications and their severity are judged by a blinded investigator who is independent of the patient’s clinical care team. The patients will be followed up at each institution for at least 90 days after surgery.

### Sample size

In an RCT involving 168 patients who underwent laparoscopic gastrointestinal surgery, the incidence of POD was 10.7% in the methylprednisolone group and 23.8% in the placebo group (16). In the pilot study conducted at the NCGG, the incidence of POD was 39.4% after gastrointestinal surgery. We hypothesize that preoperative HC treatment will reduce the incidence of POD by at least half (from 39.4% to 19.7%). Assuming an 80% power to detect an absolute difference with a two-sided alpha risk of 5%, the required sample size is 168 patients. To account for potential sample errors, we estimate that the final sample size required is 180 patients (90 per arm).

### Statistical analysis

The baseline characteristics of the enrolled patients will be summarized using descriptive statistics. The primary outcome analysis will include the full analysis set (i.e., the analysis population consists of all enrolled patients undergoing surgery), and the results of the per-protocol set (i.e., the analysis population consists of all patients receiving complete intervention) will be confirmed. Other efficacy outcomes will be evaluated in the full analysis set, whereas safety analyses will focus on patients receiving the study drug.

For the primary outcome, patients are considered to have experienced POD if they are rated as ‘positive’ at least once in the CAM-ICU test performed twice daily on postoperative Days 1–3. Conversely, they are classified as having ‘no POD’ if they are not rated as ‘positive’ even once. Logistic regression will be used to compare POD incidence between allocation groups, with the allocation group and CAM-ICU at admission used as explanatory variables.

For the secondary outcomes, continuous variables, including the change in blood sodium levels, will be compared between allocation groups. A linear mixed-effects model will be used with the interaction term between the treatment group and the time point as a fixed effect. Adjusted means and 95% confidence intervals at each time point for each treatment group will be estimated, and comparisons will be performed with multiplicity adjusted using the TukeyLKramer method.

Postoperative mortality within 90 days will be compared using Fisher’s exact test. Length of hospital stay will be analysed via the KaplanLMeier method and compared by the log-rank test. Discharge alive will be considered the event, whereas death during hospitalization will be censored at the time of death. Changes in FIM will be compared via analysis of covariance, adjusting for the baseline FIM and allocation group. To identify predictive factors for POD, a logistic regression analysis will be conducted using eight variables, including sex and seven other factors previously reported as risk factors for POD as explanatory variables: advanced age (25), low MMSE-J score (26), low preoperative serum sodium level (19), malignancy (27), regular use of benzodiazepines (28), long operation time (19), and heavy blood loss (19).

The number of patients with each perioperative adverse event/surgery-related complication and those with Grade IIIa or higher adverse events/complications will be compared using Fisher’s exact test or the χ2 test. The proportion of adverse events potentially caused by hydrocortisone will be compared using Fisher’s exact test. No interim analysis is planned.

### Unblinding

In cases in which a severe allergy, such as systemic skin rash or marked hypotension, or perioperative adverse events or surgery-related complications of ClavienLDindo classification 4a or higher occurs due to the administration of the study drug, drug administration should be discontinued immediately, and the necessary measures should be taken. In addition, open-label pharmacists should be unblinded. If the patients received the placebo, the causal agent was not HC but another postoperative drug. This unblinding protocol will allow the anaesthesiologists to properly treat severe allergies during anaesthesia. This unblinding is for the members of the independent DSMB and the principal investigator to properly evaluate the safety of HC, ensure the safety of the study, and report the SAEs to the Pharmaceuticals and Medical Devices Agency (PMDA) and the Ministry of Health, Labour and Welfare of Japan.

### Individual and entire stopping rules

If a severe allergy, such as a generalized skin rash or marked hypotension, occurs immediately after HC or placebo administration, the study will be discontinued for the patient. If surgery is cancelled (postponed indefinitely) after completion of enrolment and assignment to the study, the study will be terminated for the relevant patient. If surgery is cancelled intraoperatively due to distant metastasis or local overgrowth of the malignancy, the study will continue, and data will be collected. If surgery is postponed, the study will continue as originally assigned, with another preoperative blood test performed 7 to 1 days prior to surgery.

Discontinuation of the entire study will be considered when SAEs of Clavien–Dindo classification 4a or higher occur during the procedure through postoperative Day 28. Specifically, this will occur if there are 3 more patients requiring intensive care management or 2 more deaths in the HC group than in the placebo group.

### Trial management

The data centre at Nagoya University Hospital is responsible for the allocation sequence, EDC design and setup, web-based online enrolment and randomization. Data from the enrolled patients are anonymized and entered into the EDC. Only the investigators, members of the data centre and site monitors have access to the EDC. The anonymized data will be retrieved from the EDC after the trial. The data and research documents will be archived for 5 years after study completion. Each institution has at least one site monitor. The site monitors have the responsibility of verifying patient eligibility, obtaining written informed consent, ensuring compliance with the protocol, ensuring the accuracy of the data in the EDC, and reporting SAEs via onsite surveillance of source documents. The site monitors verify the written informed consent of all participants. For other items, the first 5 participants are verified. If there is no problem, verification is carried out every five participants thereafter. If the site monitors find any errors or deficiencies, they present these to the study team member, describe them in the monitoring report, and implement corrective actions. Auditing is not planned in this trial.

### Potential harm and reporting rules

Data on adverse events and complications are collected via the EDC. SAEs are reported to the principal investigator by the chief investigator at each institution as soon as the SAE occurs. The principal investigator must report the occurrence of SAEs to the PMDA and the Ministry of Health, Labour and Welfare of Japan using a specific reporting form. The DSMB will consider stopping the entire study if the incidence of SAEs (e.g., severe allergies, such as systemic skin rash or marked hypotension) is obviously greater in the HC group than in the other groups.

### Loss to follow-up and missing data

In general, patients participating in the current study are followed in the outpatient clinic after discharge. If the hospital visit is completed within 90 days postoperatively, the outcome is confirmed by telephone or other means. Therefore, patients can be followed up to 90 days postoperatively at each institution. In the event of missing, rejected, or abnormal data, only available data will be used to perform the analysis.

### Patient and public involvement

Patients and/or the public were not involved in the design or conduct of this study. We intend to actively involve patients and the public in the study dissemination strategy.

## Discussion

The HC-POD trial will assess whether the preoperative use of HC in digestive surgery will result in reduced POD without an increased risk of adverse events.

The presence of POD from postoperative Days 1 to 3 was selected as the primary outcome because the stress of surgery triggers POD, and HC is hypothesized to reduce POD by reducing surgical stress. To our knowledge, this trial will be the first to answer the clinical question of whether the preoperative use of HC in digestive surgery reduces POD. At study completion, we will better understand the benefits and risks of systemic HC administration in digestive surgery. We expect our findings to have broad applicability, as this trial has wide inclusion criteria and narrow exclusion criteria. We hope that this trial will have a great impact not only on the establishment of preoperative use of HC in digestive surgery but also on opening the door for preoperative HC use in other surgeries.

### Ethics and Dissemination

This protocol is based on the version (Ver.3.1) established on 19 August 2024. The study protocol was approved by the Nagoya University Clinical Research Review Board. All substantial protocol modifications will be reviewed and approved. Approval for the conduct of this study was obtained from the head of the implementing institution (Nagoya University Hospital, NCGG, Tsushima City Hospital). Investigators will obtain written informed consent from all participants. The consent forms include additional consent provisions for the collection and use of participant data for future ancillary studies (see Supplemental Additional File 2). The first patient was enrolled on 27 June 2024. To date (3 May 2025), 61 patients have been enrolled and successfully randomized. No SAEs have occurred, and unblinding has never been requested. The expected completion of patient enrolment is March 2028, and data collection will be completed in June 2028. Nagoya University Hospital is undertaking randomization, data management, statistical analysis, central trial management, and coordination. The final results of this trial will be disseminated through peer-reviewed journals and conference presentations.

## Acknowledgements

We thank all the surgeons, anaesthesiologists, open-label pharmacists, and clinical research coordinators for their efforts in conducting the HC-POD trial. The open-label pharmacists are as follows: Hiroyuki Mizoguchi, Hirotake Hida (Nagoya University Hospital); Takanobu Ichino, Hiroaki Togami, Mizuho Morishita, Humihiro Mizogami, Husa Kabumoto, and Miki Umemura (NCGG); and Shinji Ito, Junichi Isogai, Keisuke Ishikawa, Ayana Umemura, and Yuko Ito (Tsushima City Hospital).

## Authors’ contributions

T.A. and Y. Yokoyama conceived and designed this trial and were responsible for the drafting and submission of the manuscript. T.E. had a major influence on the design of this study and helped in reviewing and revising the manuscript for intellectual content. All the authors read and approved the final manuscript.

## Data availability statement

After the publication of the main article of this trial, the data will be available upon reasonable request. The data that will be shared are individual participant data that underlie the results reported in the main article after deidentification. Additionally, the study protocol and informed consent form will be available. Data will be available beginning 3 months and ending 5 years following publication of the main article for researchers who provide a methodologically sound proposal. The proposals should be sent to the corresponding author and will be reviewed by the corresponding author and Nagoya University Hospital. After approval, the data will be available in our university’s data warehouse.

