## supplemental additional file 2 for "Hydrocortisone for the prevention of postoperative delirium in digestive surgery (HyPOD): Study protocol for a multicentre randomized, blind, placebo-controlled trial"

Participant Consent Form

Principal investigator: Nagoya University Graduate School of Medicine Perioperative Medicine

Professor Yukihiro Yokoyama

I received the explanation about a randomized controlled trial “hydrocortisone to prevent postoperative delirium in patients undergoing digestive surgery” from Dr , and understood the following items. (Please check the boxes for the items you understand.)

□ Purposes, significance, methods, and expected risks of this study

□ Do not be detrimental to future medical treatment without participating in this study

□ Even if you agree once, you can withdraw your consent at any time

□ Consideration should be given to the protection of personal information

□ Method of reporting analysis results, handling of intellectual property rights, compensation

for health damage by this study

Regarding this, I agree to participate in this study under the following conditions.

Information provided in this study

□ If the information I provide will be used for other studies planned or conducted in the future,

I agree to use the information again, subject to a new ethical review and approval by the

director of the executing agency.

□ Dispose of the information after the retention period.

Date

Name (Signature)

Address
